# Re-evaluation and revision of the Eating Habits Questionnaire

**DOI:** 10.1101/2023.10.06.23296639

**Authors:** Dávid Simon, Nikolett Bogár, Szilvia Dukay-Szabó, Ferenc Túry

## Abstract

The Eating Habits Questionnaire (EHQ) is a key tool in evaluating orthorexia nervosa, but its evaluation process has witnessed considerable variation, with one item notably excluded from the last phase of its development. This study undertakes a thorough re-evaluation of the English version of the EHQ, focusing on its original 35 items, within two diverse populations where English serves predominantly as a second language. Through an online survey involving 163 female models and 243 non-models, participants completed the EHQ along with the Eating Disorder Inventory (EDI) and SCOFF questionnaire. Using Confirmatory Factor Analysis, we scrutinised the factorial validity of EHQ subscales, eliminating items that did not align with the factor structure. After eliminating 17 items from the original 35-item questionnaire, the fit of the model for EHQ was acceptable. Cronbach’s alpha values indicated acceptable reliability. The EHQ problem subscale showed significant positive correlations with all EDI subscales, while all EHQ subscales demonstrated significant positive correlations with the EDI drive for thinness subscale. Comparison of the groups based on the SCOFF threshold revealed positive and significant differences across all subscales. The study examined the impact of replacing an item in the EHQ-21 during its development process, finding that it potentially influenced the resulting factor structure. A new version of EHQ (EHQ-18) was introduced supported by the analysis of the factorial and convergent validity, as well as the reliability. Furthermore, the findings suggest a potential discriminant validity of EHQ-18 in a diverse population, mostly speaking English as a second language.

## 1. Introduction

Orthorexia nervosa (ON) is a subtype of eating disorders (ED) that shows similarities to obsessive-compulsive disorder (OCD) in relation to overly healthy eating habits (Mathieu, 2005), but is not yet included in the nosological system DSM-5-TR (American Psychiatric Association, 2022). ON was first described by Bratman (1997) as an obsessive, often extreme, and physically damaging disorder, related to but different from anorexia nervosa (AN). ON is characterised by the consumption of food considered to be pure and healthy, spending an excessive amount of time purchasing the right ingredients and preparing the appropriate meal, leading to a restrictive diet and social isolation.

Although ON is not a psychopathological entity defined by DSM-5, according to Google Scholar, the term ‘orthorexia nervosa’ occurred in 3,510 articles between 2018 and 2023. Despite the lack of an exact clinical definition of ON, there are at least 13 different ON assessment tools (Brytek-Matera, Plasonja & Décamps, 2020). However, a large number of ON assessment scales does not mean an even distribution of usage. According to a systematic review by Opitz, Newman, Mellado, Robertson & Sharpe (2020), the most commonly used measurement tool was ORTO-15 (50.0%), while the second most used tool was the Eating Habits Questionnaire (EHQ, 11.8%) developed by Gleaves, Graham & Ambwani (2013). Another systematic review investigating the up-to-date diagnostic tools and prevalence of orthorexia found ORTO-15 the most frequently used measurement tool, although the authors addressed EHQ as a tool that offers promising psychometric qualities according to relevant research (Niedzielski & Kaźmierczak-Wojtaś, 2021). The authors of both systematic reviews agreed that, despite its frequent use, ORTO-15 does not have adequate psychometric properties based on research. We found that EHQ was used or quoted in 876 articles or books between 2018 and 2023 according to the Google Scholar search for the term ‘Eating habits questionnaire’, while ORTO-15 was quoted in 1,130 publications measured by the same method. This rough estimation suggest smaller difference between the usage of ORTO-15 and EHQ, compared to the results of Opitz, Newman, Mellado, Robertson & Sharpe (2020). EHQ was developed through a three-step process. In the first step, the authors reduced the original 160 items to 59 items based on the evaluation of independent experts. In the next step, the number of items was further reduced to 35 items based on three factors using exploratory factor analysis. In the final step, a confirmatory factor analysis (CFA) was performed on an independent sample that resulted in the 21 items in a three-factor version that is used commonly. The authors of the original article found good internal consistency, test-retest reliability, convergent, and discriminant validity (Gleaves et al., 2013). According to the original article, the 35 items that were used for the final selection did not contain one item (‘The way my food is prepared is important in my diet.’) that was in the final and widely used EHQ-21.

A partial re-evaluation of EHQ was conducted by Oberle, Samaghabadi & Hughes (2017). The authors found a three-factor structure using exploratory principal component analysis; however, three of the items previously loaded on the EHQ-Problems subscale were found to be loaded on the EHQ-Knowledge subscale, renamed by the authors EHQ-Behaviour. It should also be mentioned that the authors did not test their results using CFA. The EHQ has been adapted to at least five different languages, but the factor structure and the final number of items have shown significant differences. The evaluated Italian version had the same factor construction and elements as the original version (Novara, Pardini, Pastore & Mulatti, 2017). The validated Spanish version of EHQ had a similar factor structure but consisted of 20 items (Parra-Fernández et al., 2021). The validation of the French version resulted in a three-factor structure with 16 items only (Godefroy, Trinchera & Dorard, 2021). Validation of the Polish version yielded a three-factor structure, but only with 14 items (Brytek-Matera et al., 2020). An Australian validation of EHQ resulted in a four-factor structure (Halim, Dickinson, Kemps & Prichard, 2020), but the authors used principal component analysis without further CFA on an independent sample, therefore, their results should be taken with caution.

The relative instability of the factor structure of the English and the translated version of the EHQ, the item problem related to the development of EHQ-21 and the fact that English version of EHQ was not evaluated among non-native English speakers, underline the importance of the re-evaluation of EHQ. Our study aimed to re-evaluate the English version of EHQ using the original 35 items of the second step of the original validation in two different, heterogeneous populations, among fashion models and university students who were non-native English speakers.

## 2. Methods

### 2.1 Participants

This article is a component of a broader research focused on conducting a comparative analysis between female models and non-models with an international background. The survey questionnaire was completed by 196 fashion models (sample 1) and 305 women of similar age (sample 2). Participants who did not meet the specified inclusion criteria related to age, height, and BMI were excluded from the analysis.

In Sample 1, the following inclusion criteria were used: inclusion of women with a minimum of one year of modelling experience, an age range of 16 to 37 years (with 17 cases excluded due to missing data), a minimum height of 170 cm (with 3 cases excluded due to missing data), and a BMI <25 (with 4 cases excluded due to missing data). In Sample 2, only the age limit was applied, resulting in the exclusion of 30 cases. Participants who did not provide complete data for the EHQ items were also excluded (18 cases from sample 1 and 21 cases from sample 2). Another participant was excluded due to multiple missing data. Imputation was not used to avoid introducing potential bias into the evaluation. Considering the overlaps in the excluded cases, the final sample comprised 163 models and 243 non-models. The two samples were used for the analysis together, but invariance of the factor structure was tested between the two samples.

Examination of the self-declared racial distribution revealed diversity within both samples: sample 1 included 70.6% identified as white, 2.5% as Asian, 3.7% as black, and 7.9% as other; sample 2 consisted of 91.8% white, 2.9% Asian, 1.6% black, and 3.6% other. In sample 1, there was a 29.4% proportion of missing ethnicity data, while all participants in sample 2 provided information about their ethnicity.

When comparing the two groups, the mean age of the model group and the non-model group was similar, with mean of 26.0 (SD=4.7) and 25.1 (SD=5.0), respectively. However, a notable disparity was observed in mean BMI between the model group (M=18.1, SD=1.7) and the non-model group (M=22.0, SD=4.2), with the former group showing a significantly lower BMI (p<.001).

### 2.2 Measures

The survey consisted of EHQ, items from the SCOFF questionnaire, and three diagnostic subscales of the Eating Disorder Inventory (EDI) along with general sociodemographic and anthropometric questions. The language of the questionnaire was English.

The EHQ is a measurement tool for ON consisting of three dimensions: ‘knowledge’, ‘feelings’, and ‘problems’ (Gleaves et al., 2013). Our questionnaire included all 35 items that the authors used in their evaluation research in the second part of their study. Each item (e.g., ‘I am more informed than others about healthy eating.’) is scored on a four-point scale from 1 (false, not at all true) to 4 (very true).

The SCOFF questionnaire is constructed as a simple screening tool to test mainly AN and BN (Morgan, Reid & Lacey, 1999). The questionnaire consists of five questions (for example, ‘Do you feel sick because you feel uncomfortably full?’) that should be answered yes or no, related to the main characteristics of AN and BN. The SCOFF screening threshold is at least two ‘yes’ answers.

The EDI is one of the most frequently used self-rating instruments for assessing disturbed eating attitudes and behaviour and the main psychopathological symptoms found in patients with ED (Garner, Olmstead & Polivy, 1983). The three diagnostic subscales of EDI are: Drive for Thinness (DT), Bulimia (B), and Body Dissatisfaction (BD) (Nagel, Black, Leverenz & Coster, 2020). The three diagnostic subscales of EDI consist of 23 ordered category items (for example, ‘I eat sweets and carbohydrates without feeling nervous’), each scored on a six-value scale (always, usually, often, sometimes, rarely or never), scoring 0-3 (least frequent occurrences scored by 0), where higher scores represent more severe symptoms.

### 2.3 Procedures

The current analysis is part of a comparative research that focusses on fashion models. The research was carried out using an online survey. The survey was shared by non-profit fashion model organisations, international fashion model networks, and through social media platforms (Sample 1). A similar survey was distributed among university students using the snowball method (Sample 2). The survey data did not contain personal data.

### 2.4 Statistical analysis

The normality of each item was assumed if the absolute value of skewness or kurtosis was less than 2. If distribution of items were not considered as normal, the Satorra-Bentler correction was used (Satorra & Bentler, 1988).

In the initial phase of the analysis, the factor structure of the original and widely used EHQ version (EHQ-21) was examined through Confirmatory Factor Analysis (CFA), omitting the item that was newly introduced without any pretest in the final phase of development (Gleaves et al., 2013). Subsequently, each item of the 35 in the final phase that was not included in the EHQ-21 but belonging to the same subscale as the omitted item was individually tested as a replacement item, and the factor structure of the models with each replacement item was assessed via CFA.

After checking the factor structure of the originally proposed EHQ-21, the original selection procedure, starting from the original 35 items, was repeated according to Gleaves et al., eliminating items that did not fit the factor structure using CFA, modification indices and factor loadings.

The fit of the model was measured using the comparative fit index (CFI), the Tucker-Lewis index (TLI), the standardised root mean square residual (SRMR) and the root mean square error of approximation (RMSEA). For CFI and TLI, the minimum threshold of .9 was used, according to Kline (1998), and for SRMR and RMSEA, the maximum threshold of .06 (Hu & Bentler, 1999). If the fit of the model was not proper, items were removed step by step based on modification indexes and factor loadings.

Factorial validity was also tested on the two separate subsamples, assessing configural, metric, and scalar invariance. Invariance between groups was assessed by changes in fit indices from the less constrained to the more constrained model. Based on the large sample size (N>300) and relatively equal sub-sample sizes, the following thresholds were used: DCFI<-.01 or DRMSEA<.015 (Chen, 2007).

After CFA, Cronbach’s reliability was verified by Cronbach’s α for each EHQ factor for both sub-samples and EDI factors. Cronbach’s α values of 0.7–0.95 were considered acceptable (Tavakol & Dennick, 2011).

The convergent validity of EHQ with the EDI subscales was assessed using Pearson’s correlation coefficients with two-tailed significance tests.

As ON has not yet been approved by clinical criteria, discriminant validity could not be assessed. However, we assumed that respondents above the SCOFF cut-off score have higher EHQ values. The difference was tested by the t-test or, in case of unequal variance (tested by F-test) by the Welch test independently of the distribution according to the sample size greater than 100. The effect size was measured using Cohen’s d.

A significance level of p<.05 was used for all statistical tests. SPSS 23 was used for descriptive statistics, reliability analysis, and correlation analysis. Stata 14 was used for all calculations of factorial validity (CFA).

The goals of the analysis and all statistical procedures were defined before the data collection and data-driven analyses were clearly identified and discussed.

### 2.5 Ethics

Research is in accordance with the Declaration of Helsinki and was approved by the Semmelweis University Budapest Regional Research Ethics Board (No. 3/2020). All participants gave their informed consent to participate.

## 3. Results

### 3.1 Factorial validity

The skewedness of the items was between -0.97 and 1.57, while the kurtosis was between - 1.27 and 1.60.

CFA of the original EHQ-21, excluding the newly introduced item —that was not among the 35 items used by Gleaves et al. (2013) in the final phase of the development —yielded inadequate fit according to two fit indices (GFI, RMSEA). Incorporating any items from the same subscale failed to enhance the model’s fit, as shown in Table 1.

**Table 1.** Fit statistics for the CFA of the original EHQ measurement model without the item that is not in the set of original items and with replacement of the item with any of the other items in the set belonging to the same subscale (N=406)

| Model | $\chi^2$ | df | P | CFI | TLI | GFI | RMSEA | Model AIC |
| --- | --- | --- | --- | --- | --- | --- | --- | --- |
| EHQ-21 without the item not among initial 35 | 499.74 | 167 | <.001 | .91 | .90 | .87 | .07 | 18413.4 |
| EHQ-21 with item 1 | 635.76 | 186 | <.001 | .88 | .87 | .85 | .08 | 19380.5 |
| EHQ-21 with item 26 | 645.44 | 186 | <.001 | .88 | .87 | .84 | .08 | 19324.7 |
| EHQ-21 with item 27 | 585.62 | 186 | <.001 | .90 | .88 | .86 | .07 | 19144.6 |
| EHQ-21 with item 30 | 654.38 | 186 | <.001 | .89 | .87 | .85 | .08 | 19281.5 |
| EHQ-21 with item 33 | 555.18 | 186 | <.001 | .90 | .89 | .86 | .07 | 19176.3 |

The selection process, as outlined by Gleaves et al. (2013), was repeated using the 35 original items that persisted after the prior phase of the original development procedure. The initial three-factor model with the original 35 elements did not fit our data (Table 2). After removing 17 items, the fit of the model was acceptable. The correlations between the subscales were positive and significant in the range of .37 to .70 (Table 3). Multigroup analysis had been carried out comparing fashion model and non-model samples. The configural invariance model for the two separate samples showed a fit similar to that for the full sample. The metric invariance model showed smaller changes in the fit indices compared to the thresholds. The scalar invariance model showed a smaller change in RMSEA but a larger change in CFI compared to the thresholds.

**Table 2.**
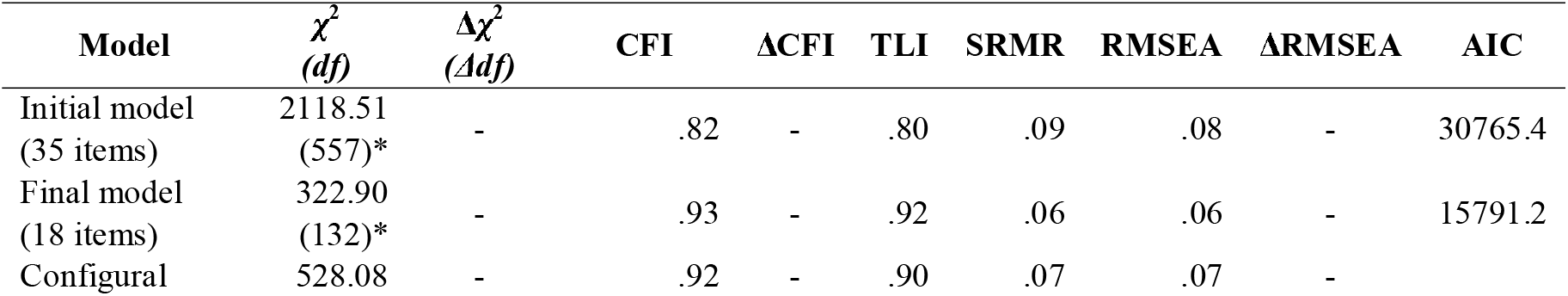
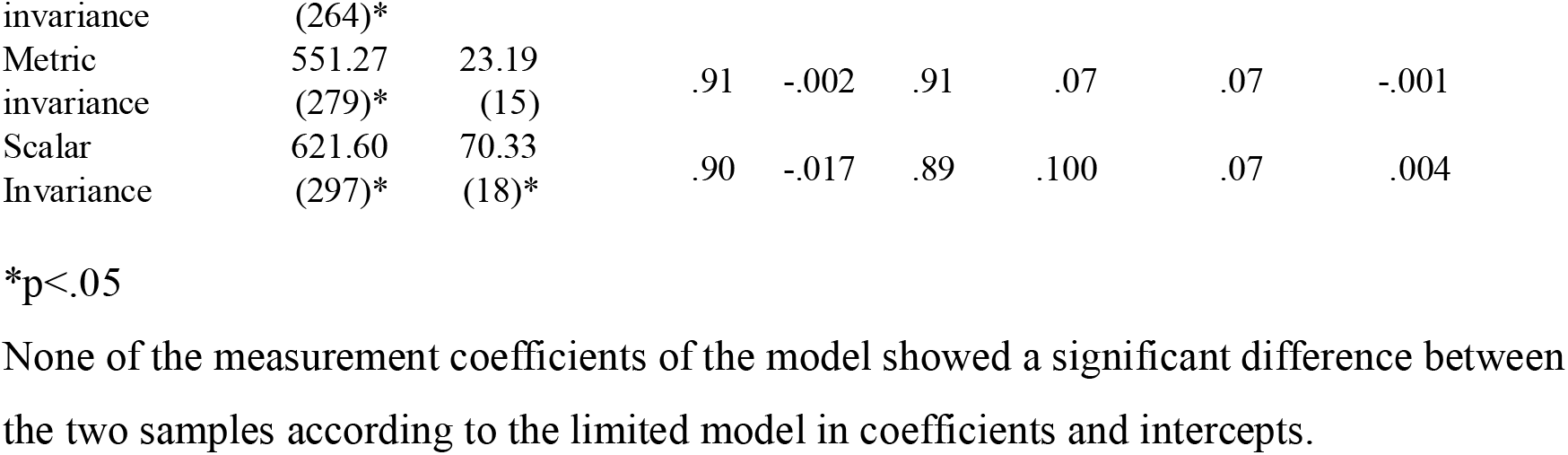
Fit statistics for CFA of the EHQ measurement models tested (N = 406)

| Model | $\chi^2$<br>(df) | $\Delta\chi^2$<br>($\Delta df$) | CFI | $\Delta CFI$ | TLI | SRMR | RMSEA | $\Delta RMSEA$ | AIC |
| --- | --- | --- | --- | --- | --- | --- | --- | --- | --- |
| Initial model<br>(35 items) | 2118.51<br>(557)* | - | .82 | - | .80 | .09 | .08 | - | 30765.4 |
| Final model<br>(18 items) | 322.90<br>(132)* | - | .93 | - | .92 | .06 | .06 | - | 15791.2 |
| Configural | 528.08 | - | .92 | - | .90 | .07 | .07 | - |  |

|  |  |  |  |  |  |  |  |  |
| --- | --- | --- | --- | --- | --- | --- | --- | --- |
| invariance | (264)* |  |  |  |  |  |  |  |
| Metric | 551.27 | 23.19 |  |  |  |  |  |  |
| invariance | (279)* | (15) | .91 | -.002 | .91 | .07 | .07 | -.001 |
| Scalar | 621.60 | 70.33 |  |  |  |  |  |  |
| Invariance | (297)* | (18)* | .90 | -.017 | .89 | .100 | .07 | .004 |

\*p<.05
None of the measurement coefficients of the model showed a significant difference between the two samples according to the limited model in coefficients and intercepts.

**Table 3.** Correlation between the EHQ and EDI subscales (N = 406)

|  | EHQ-K | EHQ-P | EHQ-F |
| --- | --- | --- | --- |
| EHQ-K | - |  |  |
| EHQ-P | .70* | - |  |
| EHQ-F | .57* | .37* | - |
| EDI-DT | .21* | .40* | .11* |
| EDI-B | .06 | .20* | .03 |
| EDI-BD | .01 | .24* | -.01 |

\* p<.05

### 3.2 Reliability

Cronbach’s alpha values for models and non-models respectively in the case of EHQ-K were .81 and .79, in the case of EHQ-P they were .90 and .87, while in the case of EHQ-F they were .80 and .77. All alpha values were in the acceptable range.

### 3.3 Convergent validity

All EHQ subscales showed a significant positive correlation with EDI-DT and EHQ-P showed a significant positive correlation with all EDI subscales, but EHQ-K and EHQ-F were not significantly correlated with EDI-B and EDI-BD (Table 3).

### 3.4 The relation of EHQ to the SCOFF result

As a simulation of discriminant validity, we measured the difference of each EHQ subscale between those who scored equal to or above the SCOFF cutoff score and the rest of the sample. The differences were positive and significant on all subscales, the effect sizes were between .4–1.2. The largest effect size was measured in the case of EHQ-P (Table 4).

**Table 4.** Differences in the means of the EHQ subscales in relation to the SCOFF cutoff score.

|  | SCOFF<2 |  |  | SCOFF≥2 |  |  | t | df | p | Cohen's d |
| --- | --- | --- | --- | --- | --- | --- | --- | --- | --- | --- |
|  | n | M | SD | n | M | SD |  |  |  |  |
| EHQ – K | 221 | 10.2 | 3.1 | 185 | 12.2 | 3.4 | -6.43 | 404 | <.001 | 0.6 |
| EHQ – P | 221 | 13.0 | 4.1<sup>+</sup> | 185 | 18.7 | 6.4<sup>+</sup> | -10.53<sup>++</sup> | 299.5<sup>++</sup> | <.001 | 1.1 |
| EHQ – F | 221 | 11.8 | 2.6 | 185 | 12.8 | 2.4 | -3.86 | 404 | <.001 | 0.4 |

<sup>+</sup> Significant difference in SD assessed by F-test.<sup>++</sup> Welch test was used according to the significant difference in SD assessed by F-test.

## 4. Discussion

The objective of the present study was to repeat the last phase of the development and the evaluation of EHQ among mainly non-native English speakers, based on the original set of items before the last phase of the original development procedure.

In the first step of the analysis, the factor structure of the widely used EHQ-21 was investigated using CFA without the item that was newly introduced in the last phase of the original development process. This factor structure was not fitted to the data. In the subsequent steps of the analysis, the missing item was replaced by each of the items from the original pool on the same subscale that were not used in the final model. None of the models with replacement items fit the data.

In the next step of the analysis, the last phase of the original development process was repeated with the original set of 35 items to find the structure of the items consistent with the whole procedure.

According to the results of the process, a three-factor EHQ solution was found and validated consisting of 18 elements (EHQ-18) on a factor structure similar to that proposed by Gleaves et al. (2013). The fit indices were similar to the original model of Gleaves et al. (2013), but our model also fitted with a restriction to equal coefficients and (less strictly) equal intercepts for the two independent samples. Consequently, configural and metric invariance was proved fully for the two independent samples, while scalar invariance was proved partially.

The differences between the elements of the currently proposed EHQ-18 version and the widely used EHQ-21 version are shown in Table 5. Regarding EHQ-K, both the original and the revised version consist of five items; however, one of the items is different: the item ‘My diet is better than the diets of other people’ was replaced by the more general and not comparable item ‘I eat only healthy foods’. item in the revised version. Similarly, only one item is different in the EHQ-F subscale: the item ‘Eating the way I do gives me a sense of satisfaction’ was replaced by the semantically similar ‘Eating healthy brings me fulfilment’. in the revised version. In contrast, the composition of the EHQ-P subscale shows more dissimilarities compared to the original version. In the revised version, the EHQ-P consists of nine items in contrast to the 13 items of the original version. More importantly, only four items are identical in the two versions. Three of the four common items (3, 5, 34) are related to social relationships, while the fourth (16) is related to the possibly compulsive character of ON. Parallelly, four of the five newly included items (2, 17, 23, 28) are related to compulsivity, while the fifth is related to the restriction of social relations. The nine items left out show less consistency. While five items are related to compulsivity (10, 14, 20, 29, 31), the remaining four items (8, 9, 15, and one without ordinal) are not closely related to any of the dimensions mentioned above.

**Table 5.** Items from the original EHQ-21 and the evaluated EHQ-18.

| No.* | Item | EHQ-21 | EHQ-18 | Factors |
| --- | --- | --- | --- | --- |
| 11 | My diet is better than other people's diets. | + |  | Knowledge |
| 13 | I am more informed than others about healthy eating. | + | + | Knowledge |
| 18 | My eating habits are superior to others. | + | + | Knowledge |
| 27 | I eat only healthy foods. |  | + | Knowledge |
| 32 | I prepare food in the most healthful way. | + | + | Knowledge |
| 33 | It's important to me to eat healthily. | + | + | Knowledge |
| 2 | I place more and more restrictions on the foods I can eat. |  | + | Problems |
| 3 | I turn down social offers that involve eating unhealthy food. | + | + | Problems |
| 5 | My healthy eating is a significant source of stress in my relationships. | + | + | Problems |
| 8 | My diet affects the type of employment I would take. | + |  | Problems |
| 9 | I have difficulty finding restaurants that serve the foods I eat. | + |  | Problems |
| 10 | I follow a health-food diet rigidly. | + |  | Problems |
| 14 | I spend more than three hours a day thinking about healthy food. | + |  | Problems |
| 15 | Few foods are healthy for me to eat. | + |  | Problems |
| 16 | I follow a diet with many rules. | + | + | Problems |
| 17 | I think about healthy food when engaged in other activities. |  | + | Problems |
| 20 | I only eat what my diet allows. | + |  | Problems |
| 23 | I take my own food with me wherever I go. |  | + | Problems |
| 24 | I avoid going out to eat with others because of my diet. |  | + | Problems |
| 28 | Most of my free time revolves around eating healthily. |  | + | Problems |
| 29 | In the past year, friends or family members have told me that I'm overly concerned with eating healthily. | + |  | Problems |
| 31 | I am distracted by thoughts of eating healthily. | + |  | Problems |
| 34 | I go out less since I began eating healthily. | + | + | Problems |
| - | The way my food is prepared is important in my diet. | + |  | Problems |
| 6 | Eating healthily brings me fulfilment. |  | + | Feelings |
| 7 | I have made efforts to eat more healthily over time. | + | + | Feelings |
| 12 | I feel in control when I eat healthily. | + | + | Feelings |
| 19 | Eating the way I do gives me a sense of satisfaction. | + |  | Feelings |
| 22 | I feel great when I eat healthily. | + | + | Feelings |

\* Numbering of items according to the original numbering in the paper by Gleaves et al.

Acceptable Cronbach alpha values regarding all subscales for both samples were supported by the reliability of the final version of EHQ similarly to the previous validations.

Convergent validity was partially assessed and also showed some important specificity, as not all EHQ subscales were correlated with EDI diagnostic subscales. The significant positive correlation between EDI-DT and all EHQ subscales shows the possible relationship between the psychopathological background of ON and other EDs. However, the fact that only EHQ-P shows a significant positive correlation with all subscales of EDI could mean that EHQ-P possibly measures the pathological dimension of orthorexia (ON), while the other two subscales could relate to healthy orthorexia (Barthels, Barrada & Roncero, 2019; Depa, Barrada & Roncero, 2019; Zickgraf & Barrada, 2022).

The simulation of discriminant validity analysis fully supports the validity of EHQ-18. However, differences in effect size values in favour of EHQ-P support the assumption that EHQ-P might measure the pathological dimension of orthorexia (ON).

Although two independent samples were drawn for the analyses, which were supposedly different in the affectedness of the ED, they could not be considered as a random sample. Furthermore, both samples were restricted to young women, taller and thinner than the average, which encourages repeating the current evaluation in a less restricted population. A further limitation of our results is that we eliminated a large number of items during CFA, based on the modification index and factor loading, increasing the possibility of overfitting our model. However, the fit of the model for both independent sub-samples decreases the probability of such an error. It should also be mentioned as a possible limitation that most of the respondents were not native English speakers, which could cause inconsistency in the answers due to misunderstandings; however, the consistent results of our analysis seem to oppose this assumption.

During the development of the EHQ-21, an item was substituted with a new one in the final phase (Gleaves et al., 2013). By excluding this item and replacing it with any of the original items, our investigation unveiled that the addition of the new item potentially influenced the resultant factor structure. Upon replicating the final phase of the development process with the original 35 items prior to the substitution, we identified an 18-item tool (EHQ-18) structured similarly into three dimensions, albeit with only 11 items overlapping with the current EHQ-21. Our analysis affirms the factorial and convergent validity, as well as the reliability, and implies the potential discriminant validity of the EHQ-18 within a specific cohort of non-native English-speaking women. The factor structure theoretically aligns with the content analysis of its components. Furthermore, our study suggests that EHQ-K and EHQ-F could effectively measure healthy orthorexia, while EHQ-P may evaluate the pathological dimension of orthorexia. However, additional research encompassing broader demographics is imperative to validate these psychometric properties and the latter assumption.

## Data Availability

All data produced in the present study are available upon reasonable request to the authors.

## Abbreviations

AN: anorexia nervosa
BMI: body mass index
BN: bulimia nervosa
CFA: confirmatory factor analysis
DSM-5-TR: Diagnostic and Statistical Manual of Mental Disorders, Fifth Edition, Text Revision
ED: eating disorder
EDI: Eating Disorder Inventory
EDI-B: Bulimia subscale of the Eating Disorder Inventory
EDI-BD: Body Dissatisfaction subscale of the Eating Disorder Inventory
EDI-DT: Drive for Thinness subscale of the Eating Disorder Inventory
EHQ: Eating Habits Questionnaire
EHQ – F: Feeling subscale of the Eating Habits Questionnaire
EHQ – K: Knowledge subscale of the Eating Habits Questionnaire
EHQ – P: Problems subscale of the Eating Habits Questionnaire
ON: orthorexia nervosa

